# Seventy Years of Mortality Transition in India: 1950-2021

**DOI:** 10.1101/2023.03.24.23287189

**Authors:** Aalok Ranjan Chaurasia

## Abstract

Mortality in India remains high by international standards. This paper analyses mortality transition in India during the 70 years since 1950 based on the annual estimates of age-specific probabilities of death prepared by the United Nations Population Division for the period 1950-2021. The analysis reveals that characterisation of mortality transition is sensitive to the summary index of mortality used. Mortality transition in India based on the geometric mean of the age-specific probabilities of death is found to be different from that based on the life expectancy at birth. The transition in mortality based on the geometric mean of age-specific probabilities of death accelerated during 2008-2019 but decelerated when based on the life expectancy at birth. The reason is that mortality transition in younger ages has been faster than mortality transition in older ages. The analysis also reveals that there were around 4.3 excess deaths associated with the COVID-19 epidemic in the country leading to a loss of around 3.7 years in the life expectancy at birth between 2019 and 2021.

## Introduction

Mortality in India remains high by international standards. Latest estimates prepared by the United Nations Population Division suggest that the life expectancy at birth in India was 67.2 years in 2021 which implies that India ranks 178 among the 236 countries or territories for which estimates have been prepared by the United Nations Population Division (United Nations, 2022). The estimates prepared by the United Nations Population Division also reveal that the rank of India vis-à-vis 236 countries of the world in terms of life expectancy at birth has never been above 160 during the period 1950 through 2021. The rank of India has been the highest (163) across the 236 countries during the period 2015 through 2018 but the lowest (196) in 1966. Between 2018 and 2021, the rank of the country decreased rapidly from 163 in 2018 to 178 in 2021, an indication that mortality transition in India slowed down considerably during the period 2018-2021 compared to mortality transition in other countries of the world. Between 2018 and 2021, the life expectancy at birth in India decreased by almost 3.7 years, from 70.9 years in 2019 to 67.2 years in 2022 (United Nations, 2022). This decrease in the life expectancy at birth in India may be attributed to the increase in mortality due to the COVID19 pandemic. India is one of the only 23 countries of the 236 countries of the world where life expectancy at birth decreased by more than 3 years between 2018 and 2021.

According to the official life tables prepared by the Registrar General and Census Commissioner of India, based on the age-specific death rates derived from the sample registration system, the life expectancy at birth in the country was 70.0 years during the period 2016-2020 or around the year 2018 (Government of India, 2022). The life expectancy at birth varies widely within the country, across states. Among the 22 states of the country for which life tables are prepared by the Registrar General and Census Commissioner of India, the life expectancy at birth is the highest in Delhi (75.8 years) but the lowest (65.1 years) in Chhattisgarh for the period 2016-2020 (Government of India, 2022). Besides Delhi, Kerala is the only other state where the life expectancy at birth is at least 75 years according to the official sample registration system of the country. On the other hand, in 9 states of the country, the life expectancy at birth is still less than 70 years.

Although, life expectancy at birth is universally used as a single summary measure of mortality, it has limitations in analysing mortality transition. A recent study has highlighted these limitations (Modig et al, 2020). The relationship between mortality and life expectancy is essentially reciprocal but the exact connection is complicated, and becomes important when life expectancy at birth is used for analysing mortality transition (Pollard, 1982). The change in the life expectancy at birth is a weighted function of the changes in mortality at individual ages plus the interaction effects of mortality changes (Pollard, 1982). The difference in life expectancies cannot be directly translated into the difference in the relative risk of mortality because both level of mortality and the distribution of mortality over age play a role (Keyfitz, 1977; Vaupel, 1986). The implicit age standardisation in the calculation of the life expectancy at birth is construed in such a way that it raises concern about the standardisation of age and, therefore, life expectancy at birth should not be used as the measure of choice to identify risk factors of death. (Modig et al, 2020). Moreover, the life expectancy at birth reflects the mortality experience of a hypothetical population and not the mortality experience of the actual population.

There are measures other than life expectancy at birth that have been suggested as the summary index of mortality but there is disagreement on the most appropriate index to analyse mortality transition. There are advantages and drawbacks of different indexes (Spiegelman, 1955; Kitagawa, 1964). Age standardised death rate is commonly used but choosing an appropriate standard population is quite difficult. Standardisation does not eliminate the effect of the differences between the age distribution of the two populations but only held it constant (Schoen, 1970). Standardisation also gives disproportionately higher weights to older ages (Yerushalmy, 1951).

Scheon (1970) has recommended that the geometric mean of the age-specific death rates, termed as ∇, should be used as the summary index of the prevailing mortality. The rationale for opting the geometric mean to construct a summary index of prevailing mortality and important properties of the index ∇ have been discussed by Schoen (1970). However, to the best of our knowledge, geometric mean of either age-specific death rates (∇) or age-specific probabilities of death which we term as index Γ has not been used for analysing mortality transition. Unlike the life expectancy at birth which depicts mortality situation of a hypothetical population, the index ∇ or the index Γ summarises currently prevailing mortality situation.

In this paper, we analyse mortality transition in India since independence or, during 1950-2021 in terms of the trend in both life expectancy at birth and geometric mean of the age-specific probabilities of death (Γ). The paper also analyses the impact of COVID-19 pandemic on mortality transition and estimates excess deaths associated with the pandemic. The paper reveals that mortality transition in India since independence has not been consistent, and there has been considerable slowdown in mortality transition because the transition in mortality has been different in different age groups. The analysis is relevant from the health policy perspective as mortality transition reflects improvements in the quality of life of the people through improvements in their health and nutritional status (United Nations, 1973). The analysis of mortality transition also contributes to understanding the evolution of the health policy. Ideally, there should be congruence between mortality transition and evolution of health policy as health policy has a direct reflection on the level and the transition in mortality. At the same time, evolution of the health policy may be viewed as a response to the health status of the population as reflected in through the transition in mortality. It is well known that with the improvement in the health status of the population, the disease profile changes, there is a change in the pattern of causes of death and a shift in the age pattern of mortality. Evolution of the health policy, therefore, is a response to the transition in mortality resulting from the improved health status of the people (Chaurasia, 2009).

The paper is organised as follows. The next section analyses long-term trend in life expectancy at birth and in the geometric mean of age-specific probabilities of death (Γ). The analysis reveals that mortality transition in the country has not been consistent as the trend in the life expectancy at birth and in geometric mean of age-specific probabilities of death (Γ) changed at least five times. Section three analyses transition in the age-specific probabilities of death (Γ) using a polishing approach that is similar to median polish frequently used in exploratory data analysis. The analysis reveals that transition in the probabilities of death in younger ages has been different from the transition in the probabilities of death in the older ages and the slow transition in the probabilities of death at older ages appears to be the reason behind slowing the improvement in the life expectancy at birth. The fourth section of the paper estimates the number of excess deaths in the country that may be associated with the COVID-19 pandemic while the last section of the paper discusses policy and programme implications of the findings of the analysis in the context of demographic and health transition.

## Long-term Trend in Life Expectancy at Birth

The United Nations Population Division has provided, for the first time, annual estimates of annual age-specific probabilities of death for the 70 years period from 1950 through 2021 for 236 countries of the word including India. These estimates permit analysis of mortality transition in each country. Using these estimates, we have analysed the trend in the life expectancy at birth during 1950-2021 to identify periods of acceleration or deceleration or even reversal in mortality transition. We have first examined whether there was a change in the trend in the life expectancy at birth or not. If there is a change in the trend, then we have identified the time point(s) or year(s) when the trend had changed so that the period 1950-2021 can be divided into different sub-periods. We have analysed mortality transition in different sub-periods by estimating annual per cent change in both life expectancy at birth and geometric mean of the age-specific probabilities of death in each sub-period assuming that the trend is linear on the log scale in each sub-period. A comparison of annual per cent change in the life expectancy at birth and in the geometric mean of age-specific probabilities of death in different sub-periods helps in identifying periods of acceleration or deceleration or reversal in mortality transition.

The jointpoint regression analysis (Kim et al, 2000) has been used for the analysis of the long-term trend as the long-term trend in mortality may not be assumed to be uniform but varies over time. There are three steps in joinpoint regression analysis. The first is to test whether there has been a change in the trend. If there is no change in the trend, then the trend analysis can be carried out by fitting a straight line (on the log scale) and the annual per cent change may be estimated from the slope of the regression line. However, if the trend has changed, then the second step involves identifying time point(s) or joinpoint(s) when the trend has changed. Finally, the last step involves estimating the regression function with identified joinpoint(s). If there are k joinpoints, then the entire reference period is divided into k+1 time-segments or sub-periods and annual per cent change (APC) in the different time segments is different. The APC in a time-segment characterises mortality transition in that time-segment. The weighted average of APC in different time-segments gives average annual per cent change (AAPC) for the entire trend period with weights proportional to the length of different time-segments. AAPC describes the long-term trend in a better way when compared to the commonly used approach in which a single regression line (on the log scale) is fitted for the entire trend period and the average annual per cent change is calculated from the slope of the regression equation (Clegg et al., 2009). This approach best summarises the trend that varies over time (Marriot, 2010).

The number of times the trend has changed can be determined statistically. There are many methods that have been proposed for the purpose. These include permutation method (Kim et al, 2000); Bayesian Information Criterion (BIC) (Kim et al, 2009); BIC3 method (Kim and Kim, 2016); and modified BIC (Zhang and Siegmund, 2007). Determining the number of times, the trend has changed statistically is driven by the data and not by any a-priori assumption.

Let *y*_*i*_ denotes the life expectancy at birth or the geometric mean of the age-specific probabilities of death for the year *t*_*i*_ such that *t*_*1*_ < *t*_*2*_ < …. < *t*_*n*_ and *k*_*1*_ < *k*_*2*_ < …. < *k*_*j*_ are the jointpoints or years when the trend has changed. Then the long-term trend in *y*_*i*_ can be modelled as

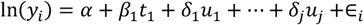

where

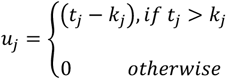

Actual calculations have been carried out using the Joinpoint Regression Program version 4.8.0.1 developed by the National Cancer Institute of the United States of America (National Institute of Health, 2020). The software requires, in advance, specification of minimum (0) and a maximum number of joinpoints (>0). The Program starts with the minimum number of joinpoints (0, which is a straight line on the Log scale) and tests whether more joinpoints are statistically significant and must be added to the model (up to the pre-specified maximum number of joinpoints). The grid search method has been used to identify joinpoints (Lerman, 1980) which allows a joinpoint to occur exactly at time *t*. A grid is created for all possible positions of the joinpoint(s) or of the combination of joinpoint(s) and then the model is fitted for each possible position of the joinpoint(s), Finally, that position of joinpoint(s) is selected which minimises the sum of squared errors (SSE) of the model. It may, however, be pointed out that even if the final selected model has *k* joinpoint(s), the slopes of all of the regression functions of the *k* + 1 time segment may not be statistically significant which means that the APC may not be statistically significantly different from zero.

Joinpoint regression analysis has been frequently used in analysing the trend in cause-specific mortality and morbidity (Akinyede and Soyemi, 2016; Chatenoud et al, 2015; Doucet et a., 2016; John and Hanke, 2015; Mogos et al, 2016; Missikpode et al, 2015; Puzo et al, 2016; Qiu et al, 2008; Rea et al, 2017; Tyczynski and Berkel, 2005). It has also been applied for estimating population parameters under changing population structure (Gillis & Edwards, 2019). Chaurasia (2020) has used it for analysing long-term trend in infant mortality rate in India. Jointpoint regression analysis is one of the methods recommended for trend analysis of health-related measures when the trend is not linear (Ingram et al, 2018). The method provides an easily interpretable characterisation of non-linear trend.

The characterisation of mortality transition in India during 1950-2021 in terms of the trend in life expectancy at birth is presented in table 1. The life expectancy at birth in India increased at an average annual per cent change (AAPC) of almost 0.69 per cent per year during 1950-2021, but the trend changed five times so that APC in different time-segments has been different. The life expectancy at birth decreased, instead increased, during 1963-66 and 2019-21, and the decrease was very marked during 2019-2021. The increase in the life expectancy at birth was relatively the most rapid during 1966-1969 but the increase slowed down subsequently and, for more than 30 years (1986-2019), life expectancy at birth in India increased at around 0.68 per cent per year. The increase in the female life expectancy at birth has been more rapid than that in the male life expectancy at birth. Moreover, the trend in male life expectancy at birth changed five times but the trend in female life expectancy at birth changed four times.

**Table 1:** Long-term trend in life expectancy at birth in India, 1950-2021.

| Segment | Endpoint |  | APC/<br>AAPC | Confidence interval |  | ‘t’ | P>\|t\| |
| --- | --- | --- | --- | --- | --- | --- | --- |
|  | Lower | Upper |  | Lower | Upper |  |  |
|  | Combined population |  |  |  |  |  |  |
| 1 | 1950 | 1963 | 0.827 | 0.791 | 0.864 | 45.099 | < 0.001 |
| 2 | 1963 | 1966 | -0.787 | -1.478 | -0.092 | -2.267 | 0.027 |
| 3 | 1966 | 1969 | 1.897 | 1.187 | 2.611 | 5.390 | < 0.001 |
| 4 | 1969 | 1986 | 1.049 | 1.022 | 1.076 | 78.034 | < 0.001 |
| 5 | 1986 | 2019 | 0.683 | 0.673 | 0.692 | 144.174 | < 0.001 |
| 6 | 2019 | 2021 | -2.687 | -3.364 | -2.005 | -7.813 | < 0.001 |
|  | 1950 | 2021 | 0.689 | 0.642 | 0.735 | 29.085 | < 0.001 |
|  | Male population |  |  |  |  |  |  |
| 1 | 1950 | 1963 | 0.773 | 0.724 | 0.822 | 31.685 | < 0.001 |
| 2 | 1963 | 1966 | -0.857 | -1.774 | 0.068 | -1.857 | 0.069 |
| 3 | 1966 | 1969 | 2.121 | 1.177 | 3.074 | 4.528 | < 0.001 |
| 4 | 1969 | 1986 | 0.881 | 0.845 | 0.917 | 49.354 | < 0.001 |
| 5 | 1986 | 2019 | 0.644 | 0.631 | 0.657 | 102.290 | < 0.001 |
| 6 | 2019 | 2021 | -2.612 | -3.512 | -1.703 | -5.709 | < 0.001 |
|  | 1950 | 2021 | 0.629 | 0.567 | 0.691 | 19.991 | < 0.001 |
|  | Female population |  |  |  |  |  |  |
| 1 | 1950 | 1962 | 0.918 | 0.841 | 0.995 | 24.013 | < 0.001 |
| 2 | 1962 | 1966 | -0.151 | -0.792 | 0.494 | -0.470 | 0.640 |
| 3 | 1966 | 1986 | 1.255 | 1.216 | 1.294 | 65.424 | < 0.001 |
| 4 | 1986 | 2019 | 0.717 | 0.700 | 0.735 | 82.034 | < 0.001 |
| 5 | 2019 | 2021 | -2.662 | -3.908 | -1.400 | -4.192 | < 0.001 |
|  | 1950 | 2021 | 0.756 | 0.702 | 0.81 | 27.623 | < 0.001 |

Source: Author
Remarks: APC – Annual per cent change
AAPC – Average annual per cent change

An assessment of mortality transition in India from the international perspective can be made by comparing the increase in the life expectancy at birth in India with the model mortality improvement trajectories developed by the United Nations (2004). These trajectories are expressed as annual increment in life expectancy at birth at a given level of life expectancy at birth but are presented as quinquennial increments. This comparison suggests that improvement in male life expectancy at birth in India has always been somewhere between medium and slow mortality trajectories (figure 1) whereas improvement in female life expectancy at birth has been close to medium trajectory (Figure 2). Mortality transition in India has always been slower than the global average.

**Figure 1:**
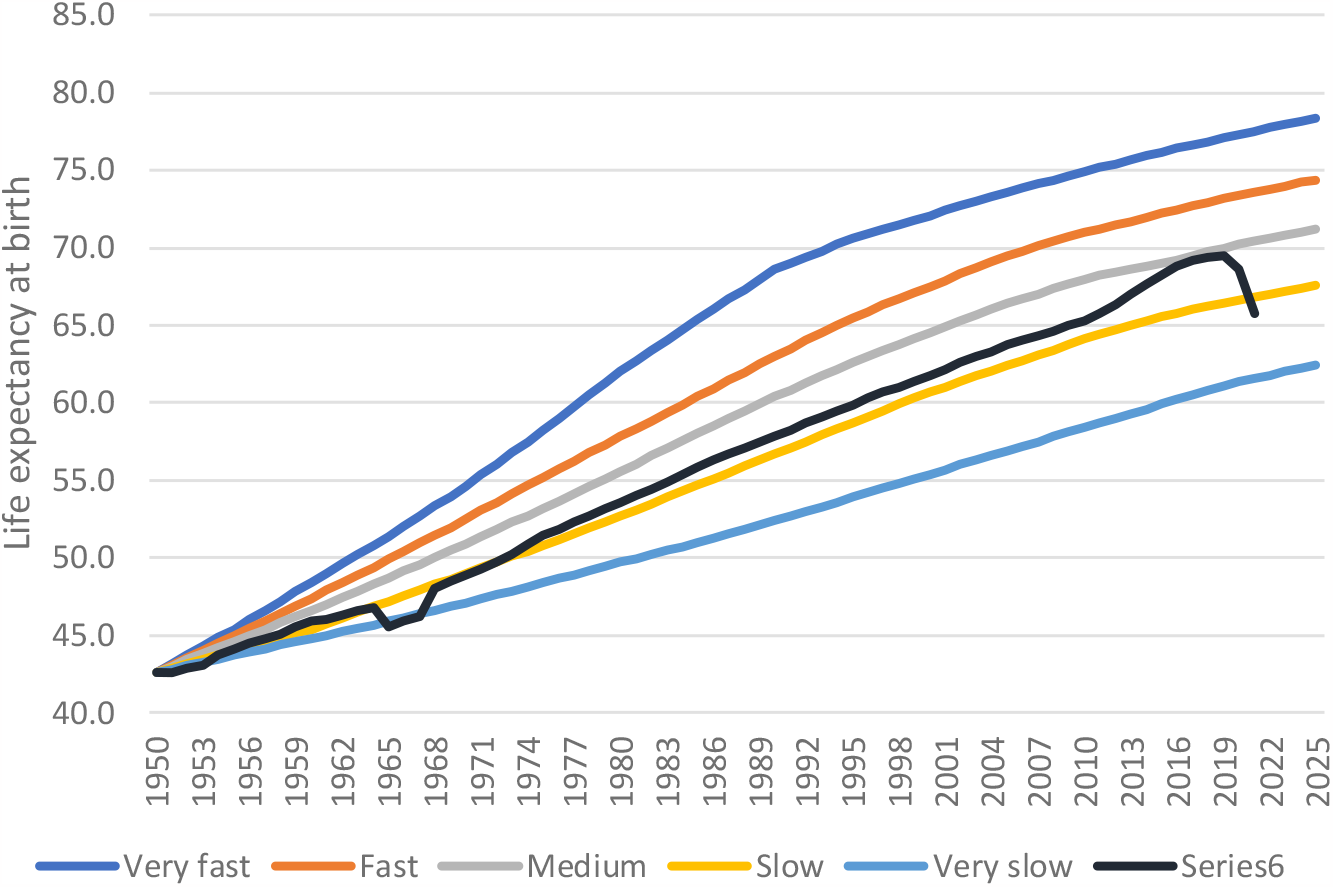
Improvement in male life expectancy at birth in India in relation to model mortality improvement schedules of United Nations

**Figure 2:**
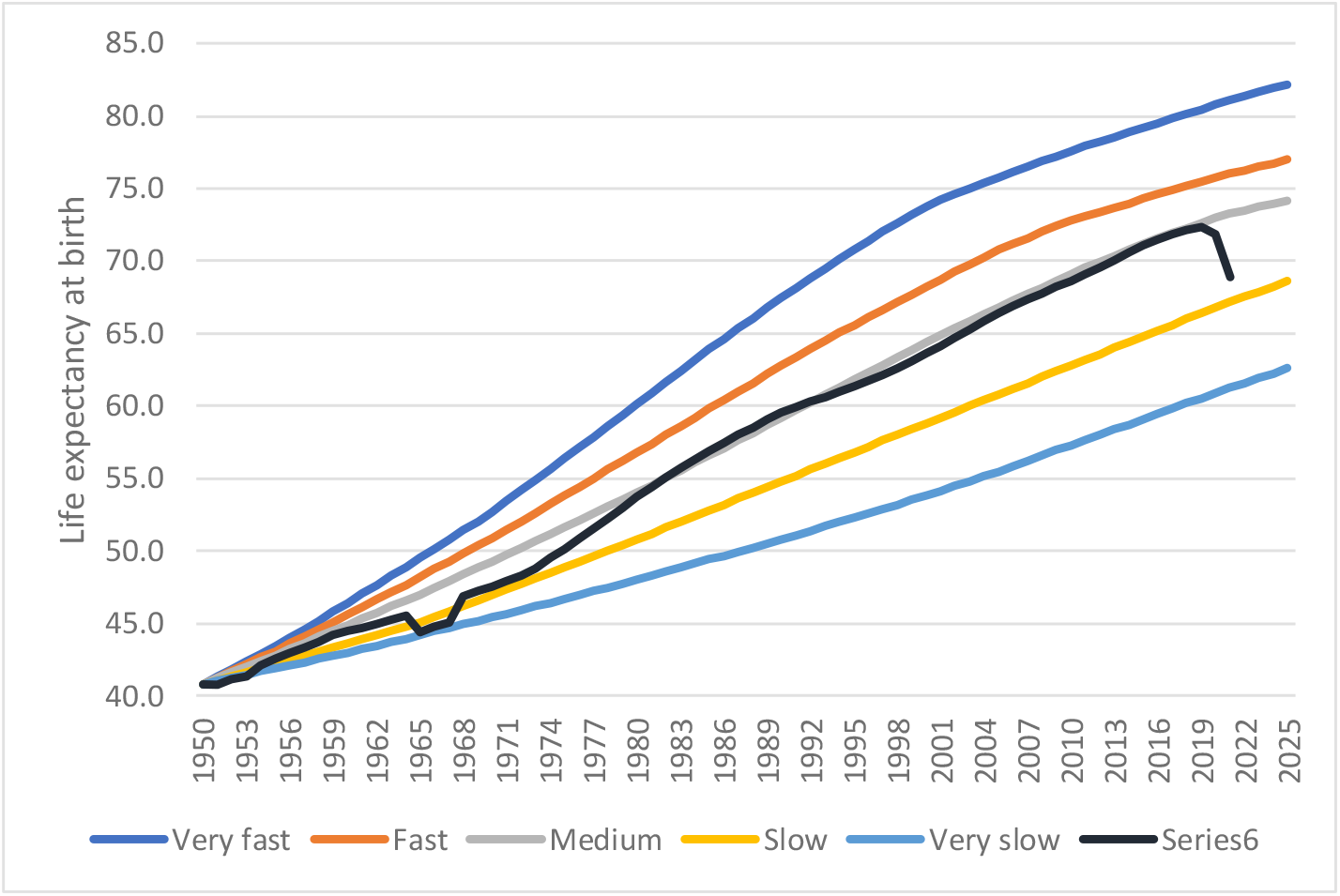
Improvement in female life expectancy at birth in India in relation to model mortality improvement schedules of United Nations

The impact of the slowdown in the improvement in the life expectancy at birth in India after 1986 appears to have been quite substantial. If the APC in the life expectancy at birth observed during the period 1969-86 would have been sustained during the period 1986-2019, the life expectancy at birth in India would have increased to more than 80 years by the year in 2019. This means that the slowdown in the life expectancy at birth in the country during the period 1986-2019 is estimated to have resulted in a loss of almost 9 years in the life expectancy at birth between 1986 and 2019. The loss in the male life expectancy at birth because of the slowdown is estimated to be almost 10 years whereas the loss in the female life expectancy at birth is estimated to have been around 8 years during the post 1986 period.

The analysis of the long-term trend in the geometric mean of the age-specific probabilities of death (Γ), however, depicts a different picture of mortality transition in the country, especially, during the period 2008-2019 (Table 2). The trend in Γ suggests that mortality transition in India has accelerated during the period 2008-2019 whereas the trend in the life expectancy at birth suggests that mortality transition has slowed down during this period. On the other hand, the trend in the life expectancy at birth suggests that mortality increased more rapidly in females compared to males during the period 2019-2021, the period of COVID-19 pandemic but the trend in the Γ suggests that mortality increased more rapidly in males as compared to that in females during the period of pandemic. Tables 1 and 2 suggest that two different summary measures of mortality depict different perspective of mortality transition in the country. The possible reason is that the life expectancy at birth gives higher weight to the probability of death in older ages whereas the index Γ, or the geometric mean of age-specific probabilities of death gives equal weight to the probability of death in all ages. This means that if transition in the probability of death in older ages is slower than the transition in the probability of death in younger ages, mortality transition depicted by the trend in the life expectancy at birth will be slower than the mortality transition depicted by the trend in the index Γ which treats transition in the probability of death in different ages equally. On the other hand, if the transition in the probability of death in older ages is faster than the transition in the probability of death in the younger ages, the trend in the life expectancy of birth will depict more rapid transition in mortality compared to the transition in mortality depicted by the index Γ. The mortality transition depicted by the trend in the life expectancy at birth will be the same as mortality transition depicted by the index Γ only when transition in the probability of death is the same in all ages. The life expectancy at birth depicts the mortality experience of a synthetic or hypothetical population whereas the index Γ depicts the mortality experience of the real population. Therefore, it is the trend in the index Γ or the geometric mean of the age-specific probabilities of death that depicts the true transition in mortality. This means that mortality transition in India has actually accelerated, not decelerated, during the period 2008-2019. Similarly, the increase in mortality during the pandemic period 2019-2021 has been more rapid in males as compared to the increase in mortality in females. Table 2 also indicates that, in the recent past, mortality transition has been faster in males as compared to the mortality transition in females.

**Table 2:** Long-term trend in the geometric mean of age-specific probabilities of death (Γ) in India, 1950-2021.

| Segment | Endpoint |  | APC/<br>AAPC | Confidence interval |  | 't' | P>\|t\| |
| --- | --- | --- | --- | --- | --- | --- | --- |
|  | Lower | Upper |  | Lower | Upper |  |  |
|  | Combined population |  |  |  |  |  |  |
| 1 | 1950 | 1962 | -0.881 | -1.021 | -0.740 | -12.505 | < 0.001 |
| 2 | 1962 | 1965 | 0.859 | -1.531 | 3.306 | 0.715 | 0.478 |
| 3 | 1965 | 1984 | -1.985 | -2.06 | -1.909 | -52.153 | < 0.001 |
| 4 | 1984 | 2008 | -1.528 | -1.580 | -1.476 | -57.898 | < 0.001 |
| 5 | 2008 | 2019 | -3.026 | -3.207 | -2.845 | -32.987 | < 0.001 |
| 6 | 2019 | 2021 | 9.981 | 7.375 | 12.65 | 7.951 | < 0.001 |
|  | 1950 | 2021 | -1.369 | -1.495 | -1.243 | -21.194 | < 0.001 |
|  | Male population |  |  |  |  |  |  |
| 1 | 1950 | 1963 | -0.759 | -0.869 | -0.648 | -13.72 | < 0.001 |
| 2 | 1963 | 1966 | 0.987 | -1.135 | 3.153 | 0.927 | 0.358 |
| 3 | 1966 | 1972 | -2.807 | -3.267 | -2.345 | -12.020 | < 0.001 |
| 4 | 1972 | 2010 | -1.365 | -1.387 | -1.342 | -119.134 | < 0.001 |
| 5 | 2010 | 2019 | -3.331 | -3.555 | -3.107 | -29.310 | < 0.001 |
| 6 | 2019 | 2021 | 10.552 | 8.229 | 12.924 | 9.469 | < 0.001 |
|  | 1950 | 2021 | -1.213 | -1.33 | -1.096 | -20.206 | < 0.001 |
|  | Female population |  |  |  |  |  |  |
| 1 | 1950 | 1963 | -0.889 | -0.997 | -0.782 | -16.528 | < 0.001 |
| 2 | 1963 | 1966 | 1.101 | -0.968 | 3.212 | 1.061 | 0.293 |
| 3 | 1966 | 1988 | -2.317 | -2.368 | -2.266 | -89.202 | < 0.001 |
| 4 | 1988 | 2000 | -1.405 | -1.543 | -1.268 | -20.353 | < 0.001 |
| 5 | 2000 | 2019 | -2.650 | -2.714 | -2.585 | -81.056 | < 0.001 |
| 6 | 2019 | 2021 | 9.321 | 7.085 | 11.604 | 8.641 | < 0.001 |
|  | 1950 | 2021 | -1.539 | -1.646 | -1.431 | -27.741 | < 0.001 |

Source: Author
Remarks: APC – Annual per cent change
AAPC – Average annual per cent change

The foregoing analysis suggests that the transition in the probability of death in the country has not been the same for all ages and it appears that transition in the probability of death in the younger ages has been faster than the transition in the probability of death in the older ages. In order to examine this hypothesis further, we have analysed the long-term trend in the age-specific probabilities of death during the period 1950-2021 under the assumption that the long-term trend in the age-specific probabilities of death has also not been linear on the Log scale during the period and the trend in the age-specific probabilities of death may have been changed at least once. The joinpoint regression analysis has therefore been carried out to analyse the long-term trend.

## Transition in Age-specific Probabilities of Death

Let *q*_*ij*_ denotes the probability of death in the year *i* and age *j*, and *q*_.._ denotes the measure of central tendency of *q*_*ij*_ over all *i* and all *j*. Then *q*_*ij*_ can be written as

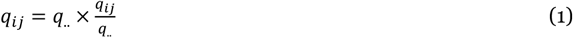

If *q*_*i*._ denotes the measure of central tendency for each *i* for all *j* and *q*_.*j*_ denotes the measure of central tendency for each *j* for all *i*, then equation (1) can be expanded as

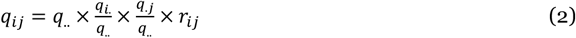

Where

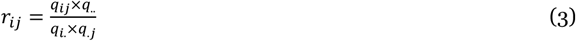

Equation (3) suggests that *q*_*ij*_ can be decomposed into four components: 1) an overall average (*q*_.._) across all *i* and all *j*; 2) a row effect (*q*_*i*._) which is common to all *j* of a given *i* and reflects how row average differs from the overall average; 3) a column effect (*q*_.*j*_) which is common to all *i* of a given *j* and reflects how column average differs from the overall average; and 4) a residual term (*r*_*ij*_) which is independent of the grand overall average, row effect and column effect.

Equation (2) can be fitted by applying the polishing technique proposed by Tukey (1977) by choosing an appropriate polishing function. The polishing algorithm successively sweeps the polishing function out of rows (divides row values by the polishing function for the row), then sweeps the polishing function out of columns (divides column values by the polishing function for the column), then rows, then columns, and so on and accumulates them in rows, columns, and in ‘all’ registers to obtain *q*_*i*._, *q*_.*j*_ and *q*_.._ and leaves behind the table of residuals (*r*_*ij*_). When the entire variation in *q*_*ij*_ across all *i* and all *j* is explained by overall average *q*_.._, row average *q*_*i*._ and column average *q*_.*j*_, all residuals (*r*_*ij*_) are equal to 1. If this is not the case, then *r*_*ij*_ reflects that part of *q*_*ij*_ which is not explained by *q*_.._, *q*_*i*._, and *q*_,*j*_. Equation (2) suggests that transition in *q*_*ij*_ should be examined after separating the overall average, the row average which is common to all ages of a given row or year in the present case and the column average which is common to all rows of a given column or age, or in terms of residuals *r*_*ij*_.

We have used Γ or the geometric mean of the age-specific probabilities of death as the polishing function because the age distribution of the probability of death is skewed. Results of the polishing exercise for the period 1950-2019 are presented in table 3 for the total population and in tables 4 and 5 for males and females respectively. The period 2020-2021 has not been included in the analysis because mortality levels during this period are biased by the COVID-19 pandemic. The overall average or *q*_.._ for all *i* and all *j* is estimated to be 0.085 for the total population, 0.087 for male population and 0.083 for female population. The polishing exercise also suggests that, in the year 1950, *q*_*i*._ or the geometric mean of the age specific probabilities of death was almost 59 per cent higher than *q*_.._ but, in the year 2019, *q*_*i*._ was almost 49 per cent lower than *q*_.._. The age pattern of average mortality across the years or the variation in *q*_.*j*_ by age is depicted in the figure 3 which shows that age pattern of average mortality in males is different from that in females.

**Table 3:**
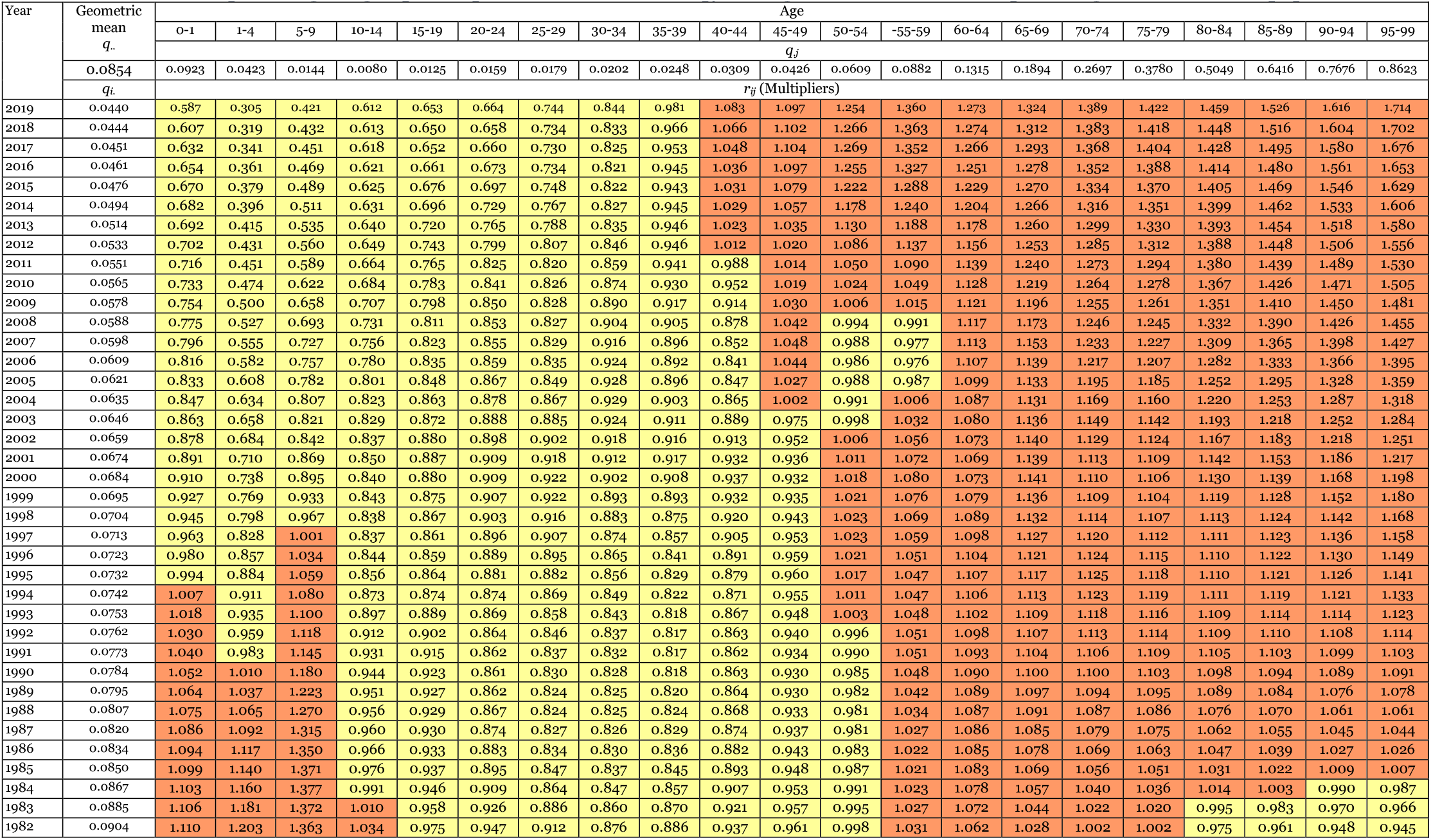
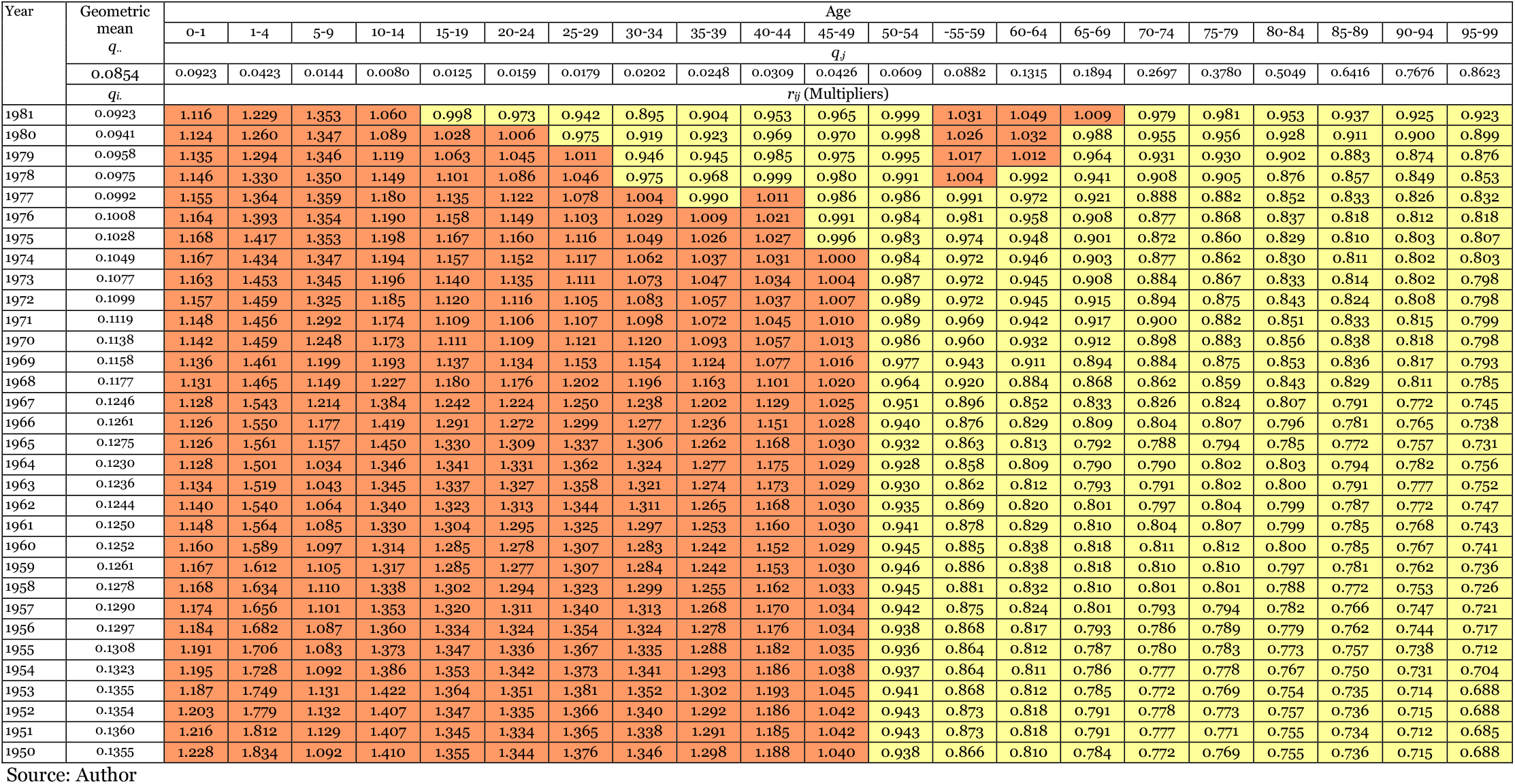
Results of the polishing of age-specific probabilities of death (*q*_*ij*_) with geometric mean as the polishing function – total population.

**Table 4:**
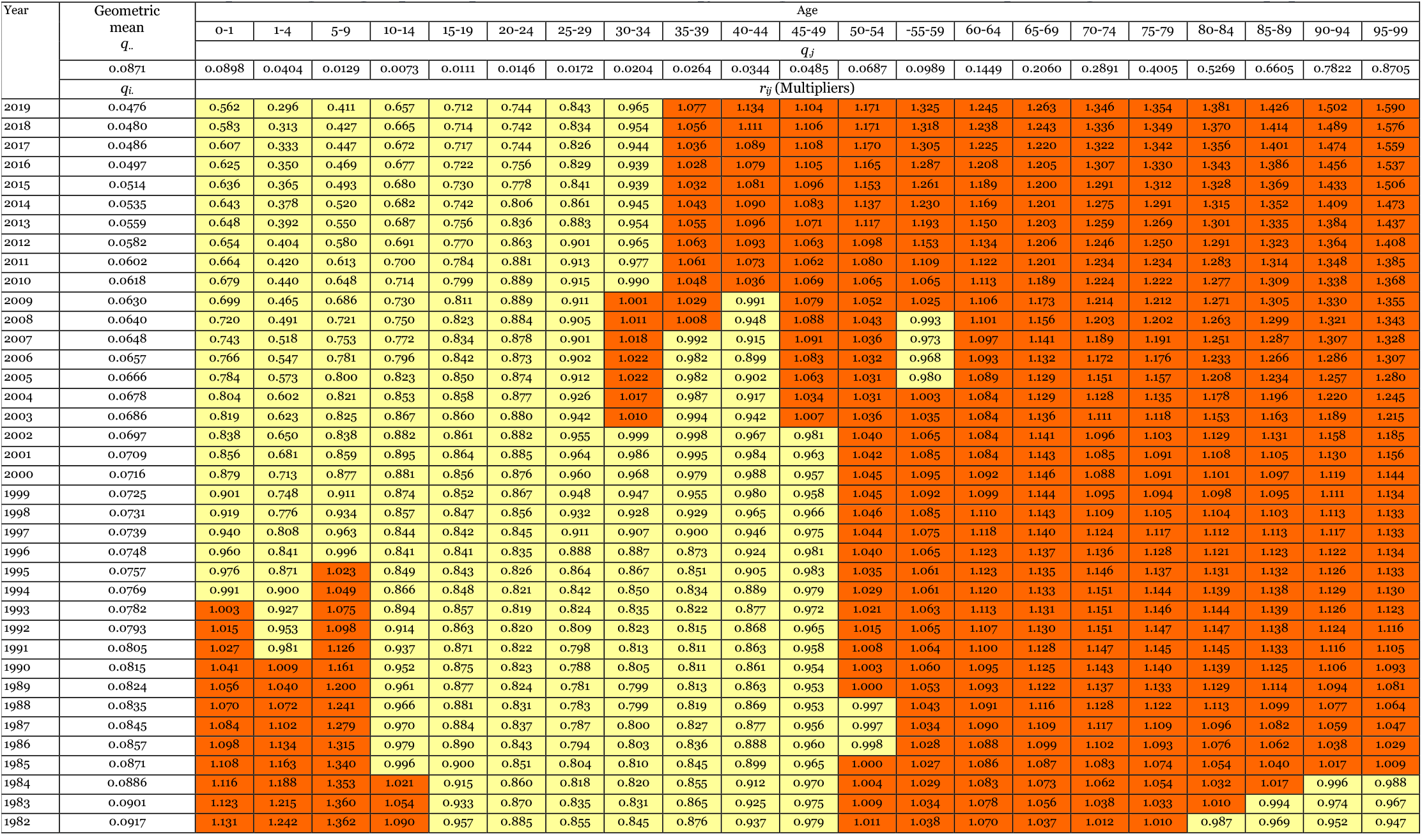
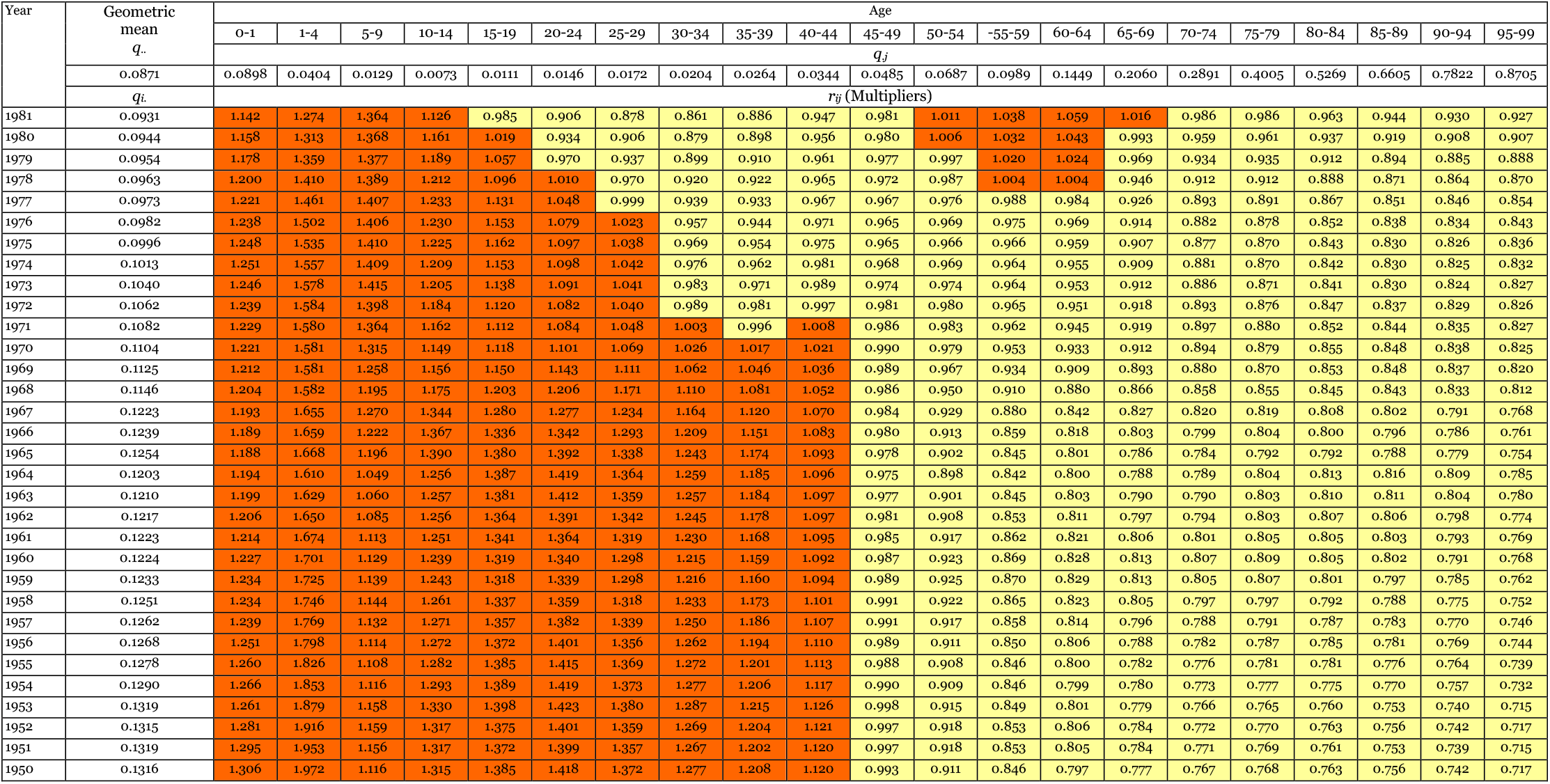
Results of the polishing of age-specific probabilities of death (*q*_*ij*_) with geometric mean as the polishing function – male population.

**Table 4:** Average annual percent change (AAPC) in *r*_*ij*_ during 1950-2019 in India

| Age | Person |  |  |  |  | Male |  |  |  |  | Female |  |  |  |  |
| --- | --- | --- | --- | --- | --- | --- | --- | --- | --- | --- | --- | --- | --- | --- | --- |
|  | AAPC % | Confidence interval |  | t | P>\|t\| | AAPC % | Confidence interval |  | t | P>\|t\| | AAPC % | Confidence interval |  | t | P>\|t\| |
|  |  | Lower | Upper |  |  |  | Lower | Upper |  |  |  | Lower | Upper |  |  |
| 0-1 | -1.066 | -1.122 | -1.010 | -37.194 | < 0.001 | -1.206 | -1.270 | -1.142 | -36.550 | < 0.001 | -0.901 | -0.932 | -0.870 | -56.215 | < 0.001 |
| 1-4 | -2.559 | -2.622 | -2.496 | -78.212 | < 0.001 | -2.694 | -2.751 | -2.637 | -90.656 | < 0.001 | -2.412 | -2.492 | -2.333 | -58.990 | < 0.001 |
| 5-9 | -1.454 | -1.564 | -1.344 | -25.720 | < 0.001 | -1.511 | -1.632 | -1.390 | -24.285 | < 0.001 | -1.359 | -1.474 | -1.243 | -22.925 | < 0.001 |
| 10-14 | -1.252 | -1.590 | -0.913 | -7.201 | < 0.001 | -0.996 | -1.243 | -0.748 | -7.844 | < 0.001 | -1.378 | -1.666 | -1.088 | -9.278 | < 0.001 |
| 15-19 | -1.098 | -1.272 | -0.925 | -12.355 | < 0.001 | -0.983 | -1.131 | -0.834 | -12.909 | < 0.001 | -1.173 | -1.391 | -0.955 | -10.483 | < 0.001 |
| 20-24 | -1.072 | -1.260 | -0.884 | -11.123 | < 0.001 | -0.961 | -1.138 | -0.783 | -10.571 | < 0.001 | -1.237 | -1.509 | -0.965 | -8.856 | < 0.001 |
| 25-29 | -0.919 | -1.050 | -0.788 | -13.666 | < 0.001 | -0.713 | -0.877 | -0.549 | -8.494 | < 0.001 | -1.151 | -1.269 | -1.033 | -19.019 | < 0.001 |
| 30-34 | -0.681 | -0.788 | -0.574 | -12.430 | < 0.001 | -0.410 | -0.565 | 0.255 | -5.159 | < 0.001 | -1.038 | -1.103 | -0.972 | -30.738 | < 0.001 |
| 35-39 | -0.412 | -0.532 | -0.292 | -6.725 | < 0.001 | -0.184 | -0.326 | -0.041 | -2.526 | 0.012 | -0.728 | -0.860 | -0.596 | -10.772 | < 0.001 |
| 40-44 | -0.146 | -0.251 | -0.042 | -2.753 | 0.006 | -0.015 | -0.108 | 0.078 | -0.314 | 0.754 | -0.326 | -0.479 | -0.172 | -4.142 | < 0.001 |
| 45-49 | 0.104 | 0.020 | 0.188 | 2.431 | 0.015 | 0.162 | 0.076 | 0.248 | 3.694 | < 0.001 | -0.016 | -0.171 | 0.138 | -0.207 | 0.836 |
| 50-54 | 0.426 | 0.302 | 0.550 | 6.736 | < 0.001 | 0.356 | 0.223 | 0.488 | 5.270 | < 0.001 | 0.519 | 0.436 | 0.602 | 12.290 | < 0.001 |
| 55-59 | 0.662 | 0.545 | 0.779 | 11.147 | < 0.001 | 0.647 | 0.519 | 0.775 | 9.945 | < 0.001 | 0.662 | 0.584 | 0.740 | 16.663 | < 0.001 |
| 60-64 | 0.666 | 0.546 | 0.786 | 10.915 | < 0.001 | 0.638 | 0.498 | 0.777 | 9.005 | < 0.001 | 0.675 | 0.544 | 0.806 | 10.150 | < 0.001 |
| 65-69 | 0.764 | 0.656 | 0.872 | 13.904 | < 0.001 | 0.679 | 0.571 | 0.787 | 12.387 | < 0.001 | 0.865 | 0.729 | 1.000 | 12.584 | < 0.001 |
| 70-74 | 0.863 | 0.759 | 0.967 | 16.369 | < 0.001 | 0.822 | 0.714 | 0.931 | 14.944 | < 0.001 | 0.935 | 0.796 | 1.074 | 13.252 | < 0.001 |
| 75-79 | 0.916 | 0.816 | 1.016 | 18.046 | < 0.001 | 0.841 | 0.744 | 0.937 | 17.110 | < 0.001 | 1.029 | 0.905 | 1.154 | 16.286 | < 0.001 |
| 80-84 | 0.967 | 0.851 | 1.083 | 16.435 | < 0.001 | 0.861 | 0.776 | 0.947 | 19.761 | < 0.001 | 1.086 | 0.966 | 1.206 | 17.813 | < 0.001 |
| 85-89 | 1.069 | 0.955 | 1.182 | 18.564 | < 0.001 | 0.925 | 0.821 | 1.030 | 17.452 | < 0.001 | 1.231 | 1.107 | 1.355 | 19.536 | < 0.001 |
| 90-94 | 1.201 | 1.094 | 1.307 | 22.196 | < 0.001 | 1.036 | 0.905 | 1.166 | 15.630 | < 0.001 | 1.367 | 1.250 | 1.484 | 23.088 | < 0.001 |
| 95-99 | 1.354 | 1.280 | 1.427 | 36.236 | < 0.001 | 1.186 | 1.117 | 1.254 | 34.162 | < 0.001 | 1.532 | 1.457 | 1.608 | 40.076 | < 0.001 |
| 100+ | 1.701 | 1.586 | 1.815 | 29.328 | < 0.001 | 1.507 | 1.404 | 1.610 | 28.873 | < 0.001 | 1.865 | 1.792 | 1.938 | 50.565 | < 0.001 |

Source: Author

**Table 5:**
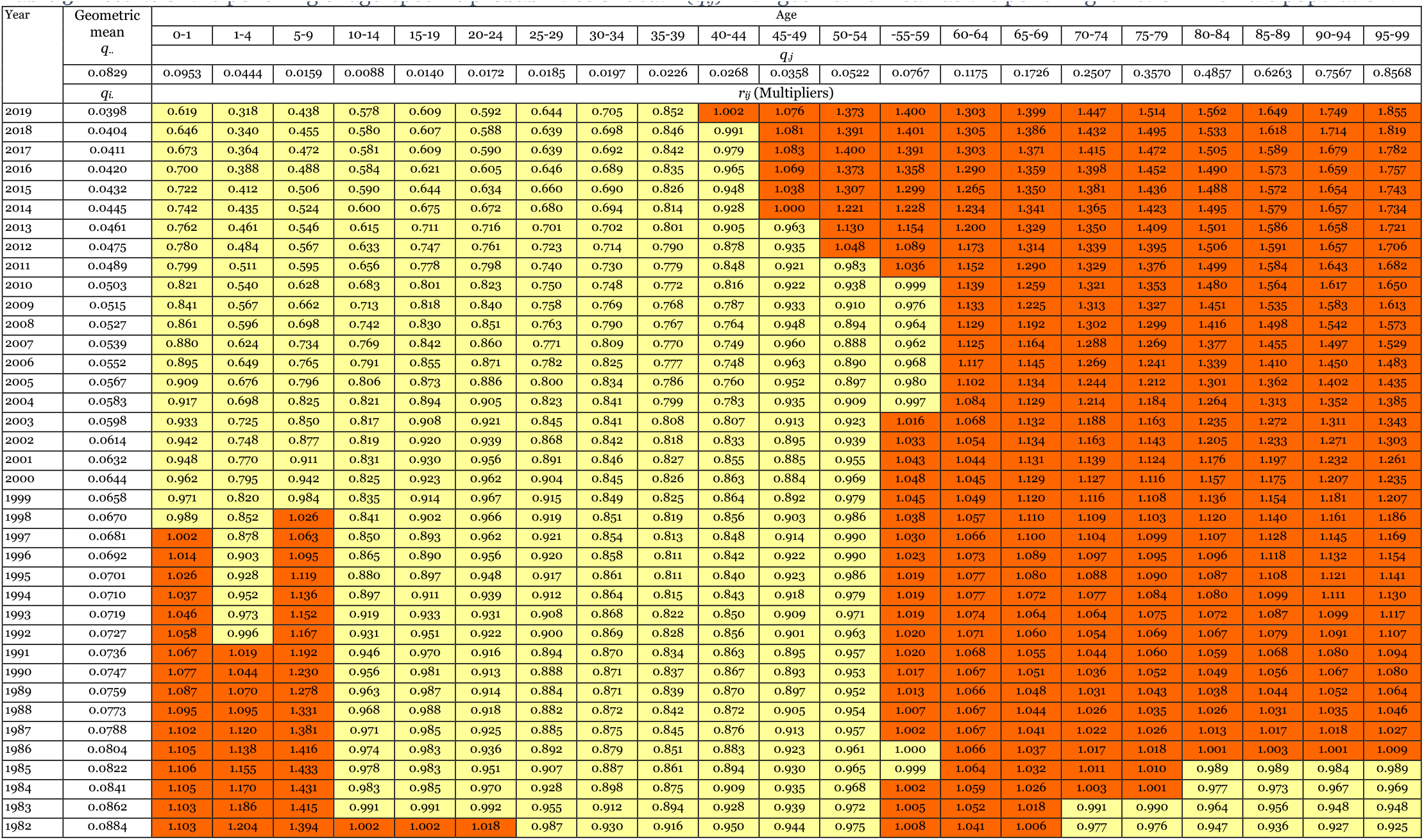
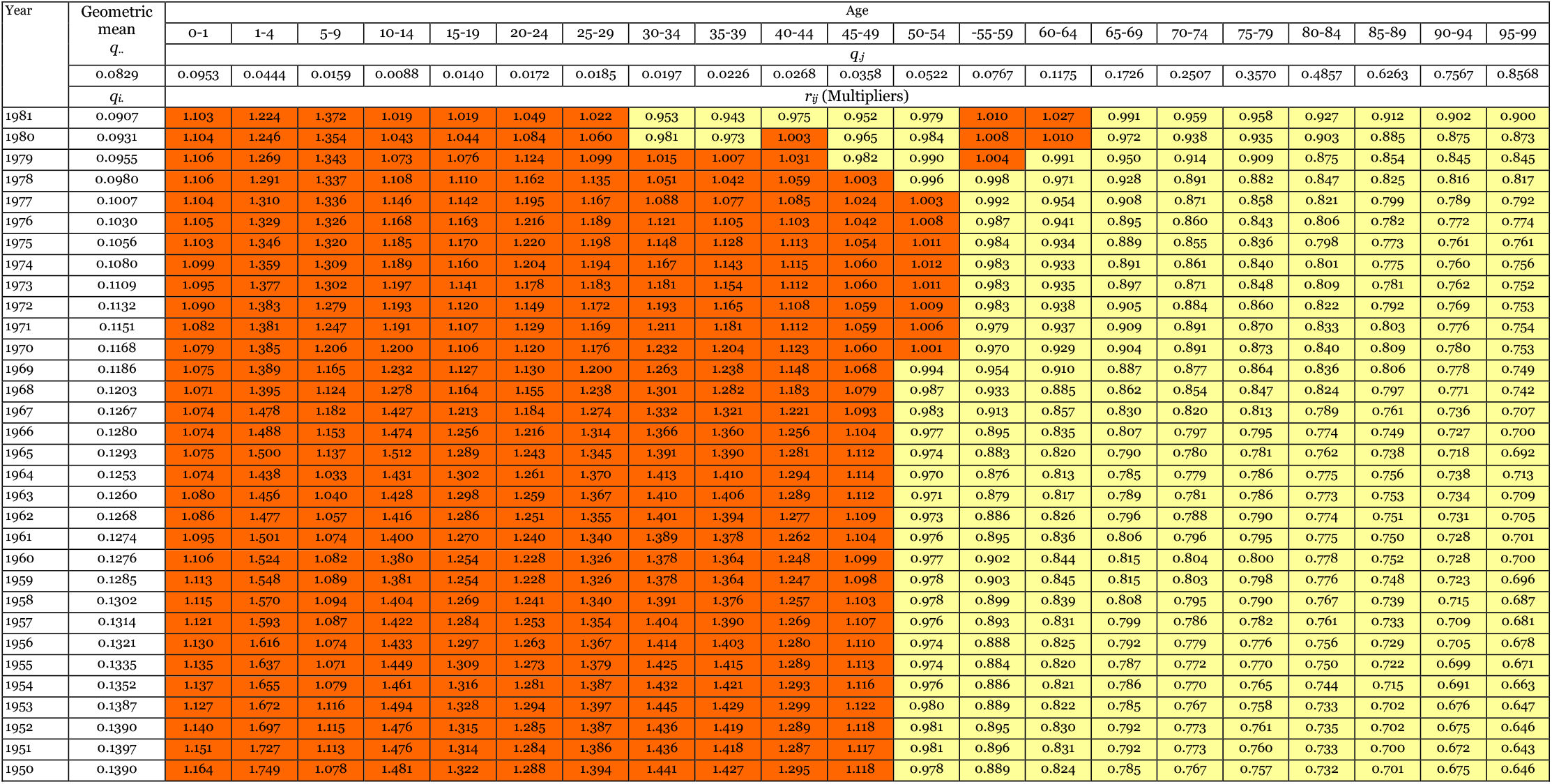
Results of the polishing of age-specific probabilities of death (*q*_*ij*_) with geometric mean as the polishing function – female population.

**Table 5:** Annual per cent change (APC) in age-specific probabilities of death in the most recent period.

| Age group | Period |  | APC | Confidence interval |  | ‘t’ | P>\|t\| |
| --- | --- | --- | --- | --- | --- | --- | --- |
|  |  |  |  | Lower | Upper |  |  |
| Male population |  |  |  |  |  |  |  |
| 0-1 | 2007 | 2019 | -4.854 | -4.937 | -4.772 | -115.453 | < 0.001 |
| 1-4 | 2006 | 2019 | -7.042 | -7.182 | -6.901 | -97.013 | < 0.001 |
| 5-9 | 2008 | 2019 | -7.987 | -8.376 | -7.597 | -39.396 | < 0.001 |
| 10-14 | 2001 | 2019 | -4.100 | -4.367 | -3.831 | -30.025 | < 0.001 |
| 15-19 | 2007 | 2019 | -4.279 | -4.607 | -3.951 | -25.607 | < 0.001 |
| 20-24 | 2017 | 2019 | -0.376 | -6.352 | 5.981 | -0.122 | 0.903 |
| 25-29 | 2010 | 2019 | -4.174 | -4.735 | -3.61 | -14.567 | < 0.001 |
| 30-34 | 2006 | 2019 | -3.522 | -3.756 | -3.287 | -29.597 | < 0.001 |
| 34-39 | 2011 | 2019 | -3.145 | -3.698 | -2.588 | -11.192 | < 0.001 |
| 40-44 | 2011 | 2019 | -2.787 | -3.411 | -2.160 | -8.815 | < 0.001 |
| 45-49 | 2007 | 2019 | -2.723 | -2.906 | -2.540 | -29.473 | < 0.001 |
| 50-54 | 2011 | 2019 | -2.047 | -2.402 | -1.691 | -11.425 | < 0.001 |
| 55-59 | 2012 | 2019 | -0.980 | -1.463 | -0.494 | -4.038 | < 0.001 |
| 60-64 | 1995 | 2019 | -1.538 | -1.594 | -1.483 | -55.164 | < 0.001 |
| 65-69 | 2016 | 2019 | 0.364 | -1.590 | 2.358 | 0.371 | 0.712 |
| 70-74 | 2009 | 2019 | -2.053 | -2.323 | -1.781 | -15.033 | < 0.001 |
| 75-79 | 2009 | 2019 | -1.921 | -2.118 | -1.723 | -19.342 | < 0.001 |
| 80-84 | 2008 | 2019 | -2.200 | -2.370 | -2.030 | -25.663 | < 0.001 |
| 85-89 | 2008 | 2019 | -2.174 | -2.336 | -2.012 | -26.572 | < 0.001 |
| 90-94 | 2008 | 2019 | -1.815 | -1.947 | -1.684 | -27.435 | < 0.001 |
| 95-99 | 2008 | 2019 | -1.392 | -1.490 | -1.293 | -28.133 | < 0.001 |
| Female population |  |  |  |  |  |  |  |
| 0-1 | 2009 | 2019 | -5.571 | -5.683 | -5.458 | -96.67 | < 0.001 |
| 1-4 | 2009 | 2019 | -8.177 | -8.394 | -7.959 | -72.22 | < 0.001 |
| 5-9 | 2000 | 2019 | -6.646 | -6.805 | -6.487 | -81.231 | < 0.001 |
| 10-14 | 2015 | 2019 | -2.368 | -4.449 | -0.242 | -2.231 | 0.03 |
| 15-19 | 2005 | 2019 | -5.610 | -5.910 | -5.309 | -36.374 | < 0.001 |
| 20-24 | 2016 | 2019 | -2.305 | -5.541 | 1.040 | -1.389 | 0.171 |
| 25-29 | 1999 | 2019 | -4.451 | -4.584 | -4.318 | -65.768 | < 0.001 |
| 30-34 | 2015 | 2019 | -1.416 | -2.634 | -0.184 | -2.303 | 0.025 |
| 34-39 | 2008 | 2019 | -1.593 | -1.868 | -1.318 | -11.512 | < 0.001 |
| 40-44 | 2006 | 2019 | -0.104 | -0.359 | 0.151 | -0.819 | 0.416 |
| 45-49 | 2011 | 2019 | -0.352 | -0.846 | 0.144 | -1.426 | 0.16 |
| 50-54 | 2016 | 2019 | -1.822 | -3.444 | -0.173 | -2.214 | 0.031 |
| 55-59 | 2008 | 2019 | 1.366 | 1.139 | 1.593 | 12.152 | < 0.001 |
| 60-64 | 2011 | 2019 | -0.949 | -1.229 | -0.668 | -6.749 | < 0.001 |
| 65-69 | 1974 | 2019 | -1.205 | -1.242 | -1.168 | -65.226 | < 0.001 |
| 70-74 | 2007 | 2019 | -1.717 | -1.881 | -1.551 | -20.695 | < 0.001 |
| 75-79 | 2012 | 2019 | -1.376 | -1.750 | -1.000 | -7.310 | < 0.001 |
| 80-84 | 2016 | 2019 | -0.053 | -1.296 | 1.206 | -0.085 | 0.933 |
| 85-89 | 2009 | 2019 | -2.240 | -2.440 | -2.040 | -22.202 | < 0.001 |
| 90-94 | 2016 | 2019 | -0.167 | -1.224 | 0.902 | -0.315 | 0.754 |
| 95-99 | 2017 | 2019 | 0.763 | -0.811 | 2.363 | 0.969 | 0.337 |

Source: Author

**Figure 3:**
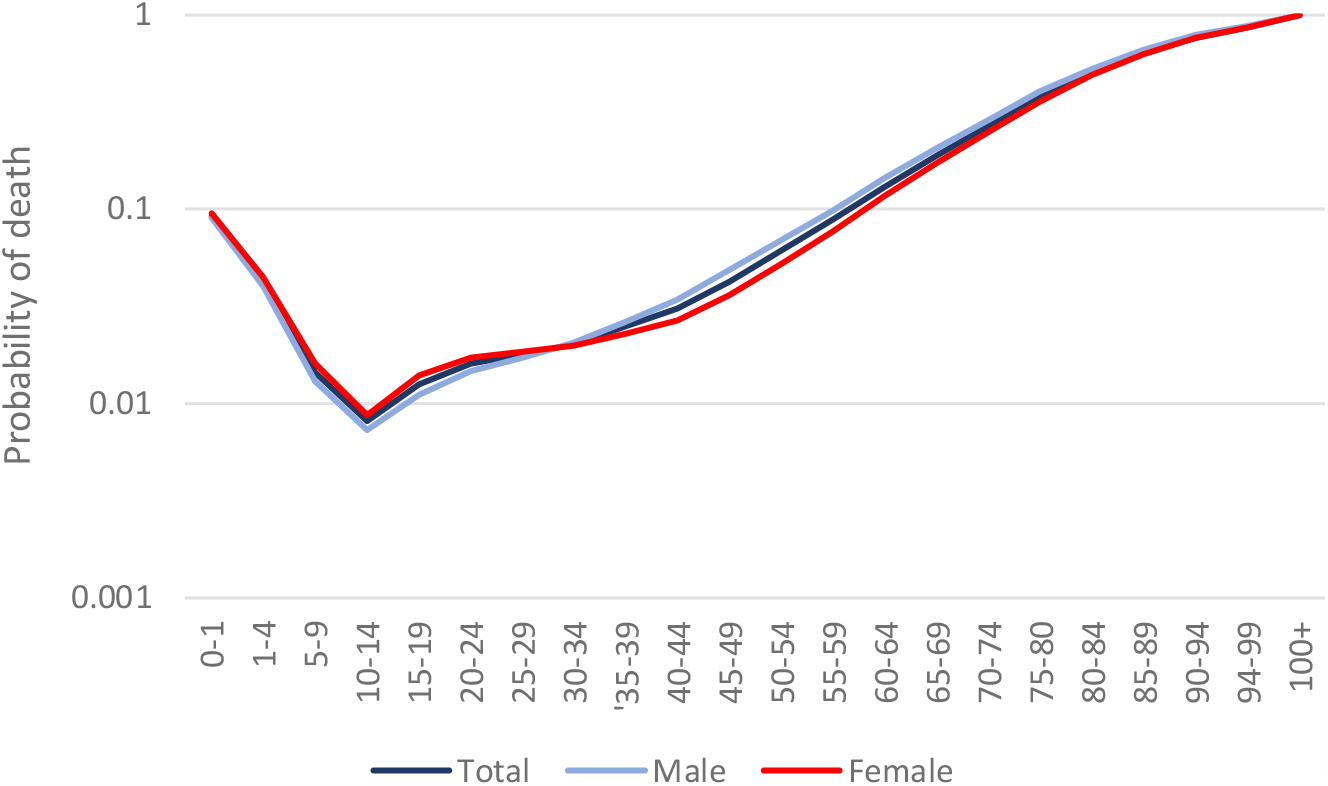
The age-pattern of average mortality during 1950-2019 in India

The residuals presented in tables 3, 4 and 5 are multipliers and their geometric mean=1. They decide whether probability of death in a particular year and age is higher or lower than the underlying probability of death determined by *q*_.._, *q*_*i*._, and *q*_.*j*_. If *r*_*ij*_>1, then the observed *q*_*ij*_ is higher than the underlying probability of death. If *r*_*ij*_<1, then the observed *q*_*ij*_ is lower than the underlying probability of death. Finally, if *r*_*ij*_=1, then the observed *q*_*ij*_ is the same as the underlying probability of death.

For example, the underlying probability of death in the age group 0-1 year determined by *q*_.._, *q*_*2019*._, and *q*_.*0-1*_ was 0.0475 in 2019, but *r*_*2019,0-1*_ was 0.5873 which means that *q*_*2019,0-1*_ was more than 41 per cent lower than the underlying probability of death determined by *q*_.._, *q*_*2019*._, and *q*_.*0-1*_. Similarly, the underlying probability of death in the age-group 70-74 years determined by *q*_*1950*._, and *q*_.*70-74*_ was 0.4277, but *r*_*1950,70-74*_ was 0.7685 so that *q*_*1950,70-74*_ was about 23 per cent lower than the underlying probability of death determined by *q*_.._, *q*_*1950*._, and *q*_.*70-74*_. The underlying probability of death in the age group 80-84 years in 2015 determined by *q*_.._, *q*_*2015*._, and *q*_.*80-84*_ was 0.212, but *r*_*2015,80-84*_ was 1.3969 so that *q*_*2015,80-84*_ was almost 40 per cent higher than the underlying probability of death determined by *q*_.._, *q*_*2015*.._, and *q*_.*80-84*_.

The transition in age-specific probabilities of death may be analysed in terms of age-specific residuals. An increase in the residuals over time is an indication that there is a deceleration in transition in the probability of death whereas a decrease in the residuals is an indication that there is an acceleration in transition in the probability of death. The time trend in residuals in different age groups is depicted in figure 4 for the total population and in figures 5 and 6 for male and female population respectively while average annual per cent change (AAPC) in total, male and female populations are presented in table 4.

**Figure 4:**
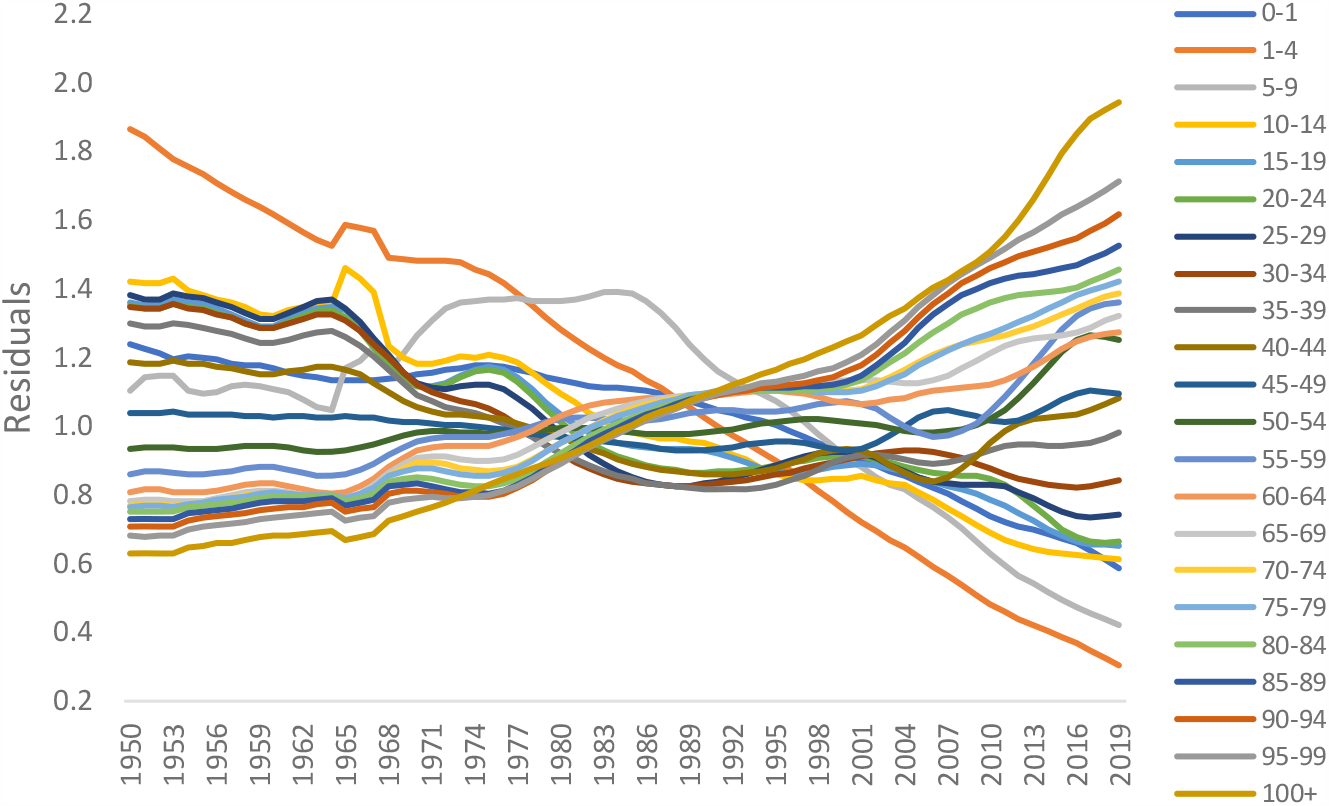
Change in age-specific residuals (r_ij_) over time – total population

**Figure 5:**
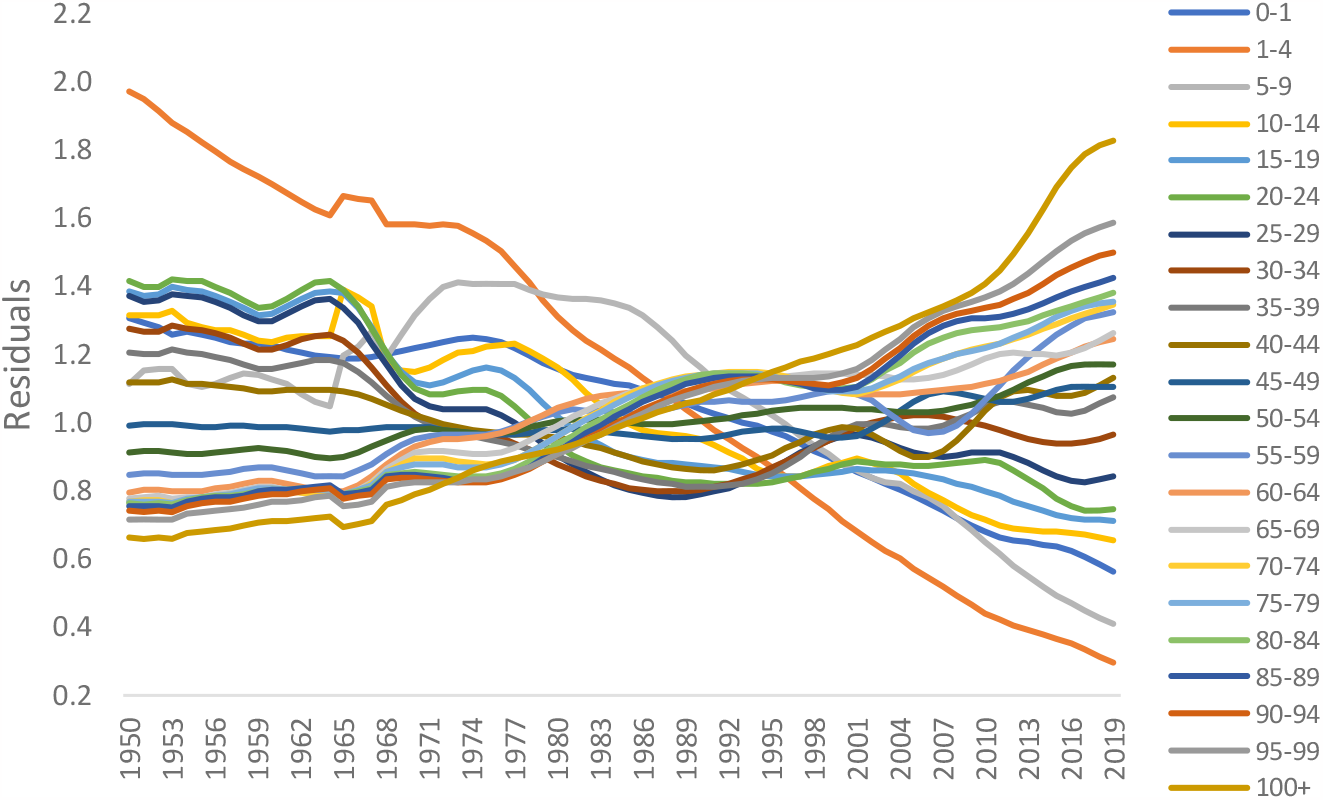
Change in age-specific residuals (r_ij_) over time - male population

**Figure 6:**
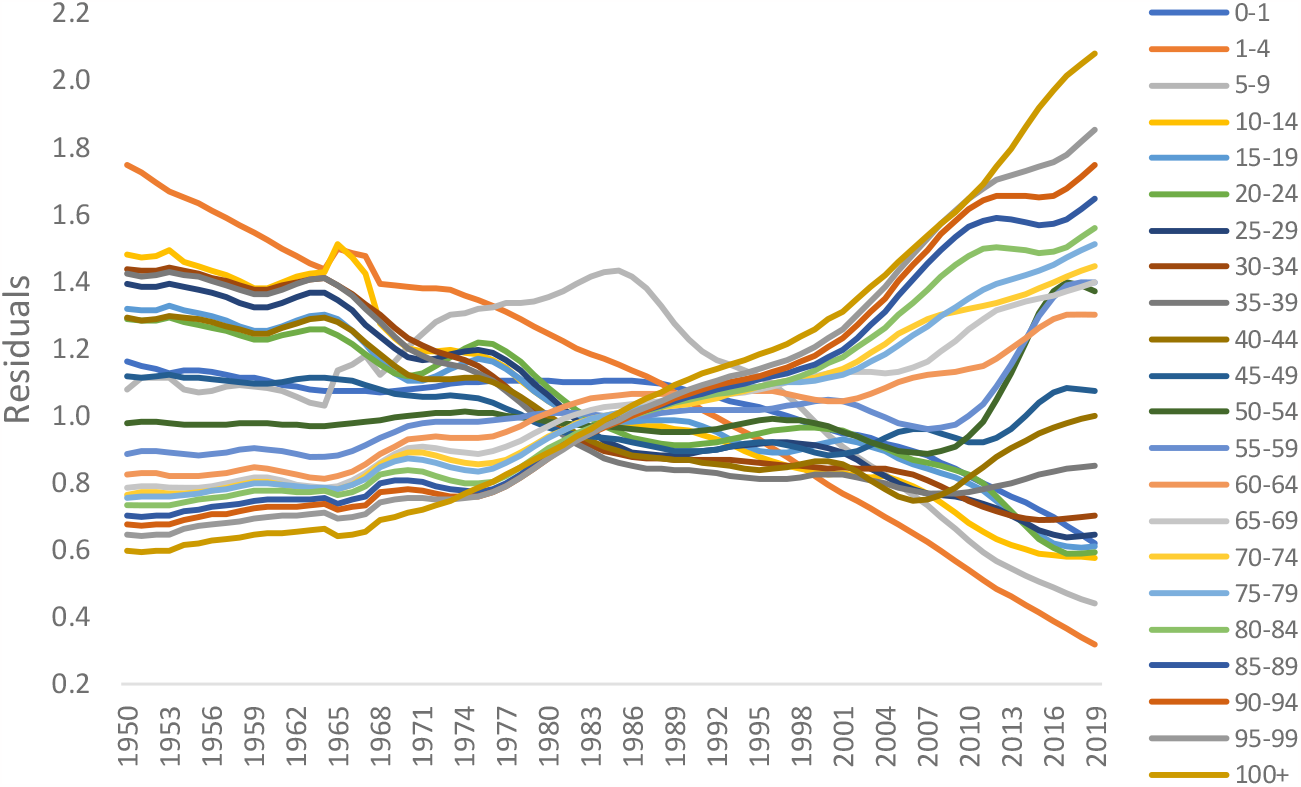
Change in age-specific residuals (r_ij_) over time - female population

Table 4 and figures 4 to 6 suggest that transition in the probability of death in different age groups in India has been different. The AAPC in *r*_*ij*_ is negative and statistically significant in ages younger than 40 years but positive and statistically significant in ages 50 years and older. This means that transition in the probability of death in ages younger than 40 years accelerated while that in ages 50 years and above decelerated over time. The transition accelerated the most rapidly in the age group 1-4 years where *r*_*i,1-4*_ decreased at a rate of more than 2.5 per cent per year during the period under reference. In 1950, *q*_*1950,1-4*_ was almost 87 per cent higher than the underlying probability of death determined by *q*_.._, *q*_*1950*,._, and *q*_.,*1-4*_ but in 2019, *q*_*1950,1-4*_ was almost 70 per cent lower than the underlying probability of death. On the other hand, there has been virtually no change in the probability of death in age groups 40-44 and 45-49 years as the AAPC in *r*_*i,40-44*_ and *r*_*i,45-49*_ has not been found to be statistically significantly different from zero.

Transition in male age-specific probabilities of death has been different than the transition in female age-specific probabilities of death. In the age group 0-10 years, transition in male probability of death accelerated more rapidly than that in female probability of death. However, in the age group 10-49 years, transition in female probability of death accelerated more rapidly than that in male probability of death. In ages 50 years and above, transition in female probability of death decelerated more rapidly than that in male probability of death. Table 4 and figures 4 to 6 suggest that transition in the probability of death in ages younger than 50 years accelerated during 1950-2021 but that in ages 50 years and older decelerated. The deceleration in transition in the probability of death in ages 50 years and older appears to responsible for the slowing down of the improvement in the life expectancy at birth.

## Impact of COVID-19 Pandemic

The estimates prepared by the United Nations suggest that the life expectancy at birth in India decreased from around 70.9 years in 2019 to 67.2 years in 2021 or a decrease of around 3.7 years. Comparable estimates of life expectancy at birth in India from the official sample registration system are not available. The decrease in the life expectancy at birth may be attributed to the excess mortality associated with the COVID-19 pandemic. It is possible to estimate excess deaths associated with COVID-19 pandemic by projecting age-specific probabilities of death that prevailed in the country in 2019 up to 2021 to obtain likely no-COVID-19 scenario and then comparing projected age-specific probabilities of death with the observed age-specific probabilities of death. The WHO defines excess mortality as, “the mortality above what would be expected based on the non-crisis mortality rate in the population of interest.” These include both direct effects of COVID-19 (deaths directly attributed to COVID-19) and the indirect knock-on effects on the health system and the society, along with deaths that are averted. (Knutson et al, 2022).

We have projected age-specific probabilities of death up to the year 2021 in the absence of COVID-19 associated mortality by estimating APC in *r*_.*j*_ for the most recent period during which the trend has been linear on the log scale for male and female population for different age groups and the results are given in table 5. The Jointpoint regression analysis was carried out to identify the most recent period in which the trend in the probability of death was linear. Based on the values of APC so obtained, we projected *r*_*ij*_ for 2020 and 2021 and then the age-specific probabilities of death in the year 2020 and the year 2021 in the absence of the deaths associated with the COVID-19 pandemic. These projected age-specific probabilities of death in the absence of COVID-19 pandemic have then been converted into the likely age-specific death rates in the absence of excess deaths associated with the COVID-19 pandemic. Based on these projected age-specific death rates in the absence of the COVID-19 pandemic, the projected number of deaths in the absence of COVID-19 pandemic have been estimated calculated for the year 2020 and 2021 for male and female population separately. The difference between the actual number of deaths in the year 2020 and 2021 and the projected number of deaths for the year 2020 and 2021 in the no-COVID-19 pandemic scenario provided an estimate of excess deaths associated with the COVID-19 pandemic in the country.

Our estimates suggest that there were around 4.29 million COVID-19 associated excess deaths – 2.44 million male deaths and 1.85 million female deaths - in India during 2020-2021. In 2020, or during the first wave of the pandemic, COVID-19 associated excess deaths are estimated to be around 0.70 million – 0.45 million male and 0.25 million female. COVID-19 associated excess deaths, however, increased to more than 3.59 million – 1.99 million male and 1.60 million female - in 2021 or during the second wave of the pandemic. The estimated number of COVID-19 associated excess deaths are presented in table 6. The number of COVID- 19 associated excess death increased rapidly after 45 years of age and peaked in the age group 65-69 years in both 2020 and in 2021. More than 60 per cent of the excess deaths associated with the COVID-19 pandemic were confined to ages above 60 years.

**Table 6:** Excess deaths (million) associated with COVID-19 pandemic.

| Age | 2020 |  |  | 2021 |  |  | 2020-2021 |  |  |
| --- | --- | --- | --- | --- | --- | --- | --- | --- | --- |
|  | Total | Male | Female | Total | Male | Female | Total | Male | Female |
| 0-1 | -0.032 | -0.016 | -0.016 | -0.051 | -0.026 | -0.025 | -0.083 | -0.042 | -0.041 |
| 1-4 | -0.010 | -0.005 | -0.005 | -0.017 | -0.009 | -0.008 | -0.027 | -0.014 | -0.014 |
| 5-9 | -0.003 | -0.002 | -0.002 | -0.006 | -0.003 | -0.003 | -0.009 | -0.004 | -0.005 |
| 10-14 | -0.002 | -0.001 | -0.001 | -0.002 | -0.001 | -0.001 | -0.004 | -0.002 | -0.002 |
| 15-19 | -0.011 | -0.007 | -0.004 | 0.018 | 0.008 | 0.010 | 0.007 | 0.001 | 0.006 |
| 20-24 | -0.017 | -0.012 | -0.005 | 0.029 | 0.013 | 0.016 | 0.012 | 0.001 | 0.011 |
| 25-29 | -0.018 | -0.013 | -0.005 | 0.035 | 0.017 | 0.018 | 0.018 | 0.005 | 0.013 |
| 30-34 | -0.013 | -0.010 | -0.003 | 0.051 | 0.030 | 0.021 | 0.038 | 0.020 | 0.018 |
| 34-39 | -0.004 | -0.003 | -0.001 | 0.076 | 0.047 | 0.029 | 0.072 | 0.044 | 0.028 |
| 40-44 | 0.009 | 0.007 | 0.002 | 0.101 | 0.063 | 0.038 | 0.110 | 0.069 | 0.041 |
| 45-49 | 0.019 | 0.014 | 0.005 | 0.156 | 0.098 | 0.057 | 0.175 | 0.112 | 0.063 |
| 50-54 | 0.041 | 0.027 | 0.014 | 0.235 | 0.136 | 0.098 | 0.276 | 0.164 | 0.112 |
| 55-59 | 0.071 | 0.048 | 0.023 | 0.312 | 0.189 | 0.123 | 0.383 | 0.237 | 0.146 |
| 60-64 | 0.112 | 0.072 | 0.040 | 0.421 | 0.236 | 0.184 | 0.532 | 0.308 | 0.224 |
| 65-69 | 0.141 | 0.087 | 0.054 | 0.490 | 0.274 | 0.216 | 0.631 | 0.361 | 0.270 |
| 70-74 | 0.126 | 0.074 | 0.051 | 0.479 | 0.260 | 0.219 | 0.604 | 0.334 | 0.270 |
| 75-79 | 0.083 | 0.051 | 0.032 | 0.409 | 0.222 | 0.187 | 0.492 | 0.273 | 0.219 |
| 80-84 | 0.091 | 0.060 | 0.031 | 0.409 | 0.212 | 0.197 | 0.501 | 0.272 | 0.228 |
| 85-89 | 0.084 | 0.057 | 0.027 | 0.316 | 0.158 | 0.158 | 0.400 | 0.215 | 0.185 |
| 90-94 | 0.028 | 0.020 | 0.008 | 0.106 | 0.054 | 0.052 | 0.134 | 0.073 | 0.061 |
| 95-99 | 0.005 | 0.004 | 0.002 | 0.022 | 0.011 | 0.011 | 0.027 | 0.015 | 0.012 |
| 100+ | 0.000 | 0.000 | 0.000 | 0.003 | 0.001 | 0.001 | 0.003 | 0.002 | 0.001 |
| Total | 0.701 | 0.454 | 0.246 | 3.591 | 1.991 | 1.601 | 4.292 | 2.445 | 1.847 |

Source: Author

There is reasonable degree of agreement between our estimates of COVID-19 associated excess deaths and estimates prepared by World Health Organization and other researchers. The World Health Organization has estimated around 4.7 million COVID-19 associated deaths in India between January 2020 and December 2021 and around 4.3 million deaths between June 2020 and June 2021 (Knutson et al, 2022). Jha et al (2022), using data collected from a nationally representative telephone survey and official data related to facility-based COVID-19 deaths and the deaths registered under the civil registration system in 10 states, have estimated more than 3.2 million COVID-19 associated excess deaths during June 2020 to June 2021. Anand et al (2021), using different approaches, estimated that COVID-19 associated excess deaths ranged from around 3.4 million to around 4.9 million during April 2021 to June 2021.

## Discussions and Conclusions

Mortality transition has not been an area interest in demographic research in India in recent years. There is no article on mortality transition in the country that has been published in Demography India, the official publication of the Indian Association for the Study of Population, since 2015. There have been studies on mortality transition in India in the past (Bhat, 1987; 1989; Bhat and Navaneetham, 1991; Chaurasia, 2006; 2009; Guha, 1991; Jain et al, 1985; Navaneetham, 1993; Preston and Bhat, 1984; Roy and Lahiri, 1987; Visaria, 2004) but, in recent years, interest in mortality transition in the country appears to have waned. The present analysis is probably and so obviously the first to analyse mortality transition in the country since independence.

A revealing finding of the present analysis is that characterisation of mortality transition is sensitive to the summary index of mortality used. Mortality transition in India based on the trend in the geometric mean of the age-specific probabilities of death (Γ) is found to be different from that based on the trend in the life expectancy at birth. The trend in the geometric mean of age-specific probabilities of death suggests that mortality transition has accelerated in the country during 2008-2019 whereas the trend in the life expectancy at birth suggests that transition has decelerated. In 2005, India launched the National Rural Health Mission which was effective from 2007, the period 2005-2007 being the preparatory phase. The Mission was directed towards providing equitable, affordable, and quality health care to the rural population of the country, especially the vulnerable population groups with especial focus on states where mortality transition was slow (Government of India, 2005). The thrust of the Mission was on establishing a fully functional, community-owned, decentralized health delivery system with inter-sectoral convergence at all levels, to ensure simultaneous action on a wide range of determinants of health including water, sanitation, education, nutrition, social and gender equality. In 2013, India launched the National Urban Health Mission to meet the health needs of the urban population with a focus on making available essential primary health care services (Government of India, 2013a). Subsequently, the two Missions were merged to constitute the National Health Mission (Government of India, 2013b). The trend in the geometric mean of age-specific probabilities of death suggests that these Missions have contributed towards accelerating mortality transition in the country whereas the trend in the life expectancy at birth indicates that there has been little impact of these Missions on mortality transition. The trend in the life expectancy at birth also implies that the rapid economic growth that India has witnessed since 2000 also had little impact in accelerating mortality transition in the country.

The contrasting scenario of mortality transition revealed by the trend in geometric mean of the age-specific probabilities of death and the trend in the life expectancy at birth appears to be due to different weights assigned to age-specific probabilities of death in the calculation of the geometric mean of the age-specific probabilities of death and in the calculation of the life expectancy at birth. In the calculation of the geometric mean, equal weights are assigned to the probability of death in different age groups. In the calculation of the life expectancy at birth, higher weights are assigned to the probability of death in older ages compared to the weights assigned to the probability of death in younger ages. As a result, transition in mortality in older ages has a stronger impact on the improvement in the life expectancy at birth. The deceleration in the life expectancy at birth reflects the deceleration in mortality transition in older ages. The present analysis reveals that mortality transition in older ages has decelerated in India during 1950-2019. From the policy and programme perspective, the present analysis suggests that the geometric mean of the age-specific probabilities of death (index Γ) or the geometric mean of the age-specific death rates (index ∇) should be preferred over the life expectancy at birth as the summary index of mortality to analyse mortality transition as the geometric mean of age-specific probabilities of death or age-specific death rates reflects the mortality experience of the real population.

A comparison of the trend in the geometric mean of the age-specific probabilities of death and the trend in the life expectancy at birth provides valuable insight about mortality transition in different age groups. If the annual per cent change (APC) in the geometric mean of the age-specific probabilities of death and the annual per cent change (APC) in the life expectancy at birth is the same, then it can be interpreted that mortality transition is the same in all age groups. If APC in the geometric mean of the age-specific probabilities of death is higher than APC in the life expectancy at birth, then it can be interpreted that mortality transition is faster in younger ages than in older ages. Finally, if the APC in the geometric mean of age-specific probabilities of death is slower than the APC in the life expectancy at birth, then this means that mortality transition is faster in older ages than in younger ages. In India, mortality transition in younger ages has been faster than that in the older ages since independence. The pace of transition has accelerated the most in the age group 1-4 years. The pace of transition decreases with age and is the slowest in the age group 40-44 years. In ages 45 years and above, the pace of transition has decelerated, and the deceleration increased with age.

The analysis also reveals interesting differences in mortality transition in males as compared to females. In younger ages (less than 10 years), mortality transition has been faster in males compared to females. However, in the age group 10-49 years, mortality transition has been faster in females compared to males. In population 50 years and older, mortality transition has been slower in females than in males as the deceleration in the probability of death in the older ages has been more pronounced in females as compared to males.

The present analysis also confirms that the impact of COVID-19 pandemic on mortality in India has been quite substantial as the pandemic appears to have resulted in a loss of almost 3.7 years in the life expectancy at birth and accounted for at least 4 million pandemic associated excess deaths in the country. The loss in the life expectancy at birth in India associated with the pandemic has been amongst the highest across countries for which estimates are prepared by the United Nations Population Division. Moreover, there have been more male excess deaths with the pandemic compared to female deaths.

The analysis suggests that the National Rural Health Mission launched in 2005 and the National Urban Health Mission launched in 2013 have contributed to accelerating mortality transition in the country. This may be viewed as the success of these Missions in meeting the health care needs of the people of the country, although there appears to have been a deceleration in mortality transition in the older ages.

The present analysis calls for promoting health in the substantive and not in the formal context. In the formal context, health of the people is linked to institution-based care whereas, in the substantive context, health is linked to the interaction between the man and the environment. There is a need to strike a balance between the two. Strengthening formal health care is related to creating, strengthening, and expanding the health care delivery system – public or private. In the absence of substantive health care, however, the system may remain unaccepted by the people at large and hence underutilised.

## Data Availability

All data produced in the present study are available upon reasonable request to the authors

